# Mitochondrial DNA haplogroup variation in hydrocephalus

**DOI:** 10.1101/2022.08.15.22278803

**Authors:** Tina N Munch, Paula L Hedley, Christian M Hagen, Joanna Elson, Marie Bækvad-Hansen, Frank Geller, Jonas Bybjerg-Grauholm, Merete Nordentoft, Anders Børglum, Preben B Mortensen, Thomas M Werge, Mads Melbye, David M Hougaard, Michael Christiansen

## Abstract

Hydrocephalus is a genetically and phenotypically heterogenous condition with complex etiology. Ciliary dysfunction has been shown to play a role, either through interference with signaling functions in primary cilia, cerebrospinal fluid flow by motile cilia, or both. Ciliary function is highly energy-dependent, consequently, variation in mitochondrial OXPHOS function might be a susceptibility factor for hydrocephalus. Furthermore, familial hydrocephalus exhibits preferential maternal inheritance. Mitochondrial DNA (mtDNA) haplogroups, have been associated with different characteristics of OXPHOS function as well as susceptibility to autism spectrum disorders, a frequent co-morbidity of hydrocephalus. This nested case-cohort study, a substudy of the iPSYCH study, used mtDNA data from 191 hydrocephalus cases and 24,831 population controls and found no association between hydrocephalus and any mtDNA haplogroup. Likewise, the distribution of European macro-haplogroups, HV, JT, and UK, did not differ between 172 hydrocephalus cases and 21,850 population controls. Thus, mtDNA haplogroups are not susceptibility factors for hydrocephalus.

## Introduction

Hydrocephalus (HC) is a heterogeneous disorder characterized by the abnormal amounts of cerebrospinal fluid (CSF) within the ventricles and subarachnoid space of the brain.[1] It is a common disorder with a frequency of around 1:1000 children.[2] Primary HC, e.g., HC without underlying tumors, hemorrhage or CNS infections obstructing or impeding the normal flow or absorption of CSF may present as syndromic, as well as isolated HC. Primary HC may be caused by congenital obstructions of CSF outlets in the ventricular system, the most common being stenosis of the Sylvian aqueduct that connects the third and fourth ventricle. Communicating forms of primary HC may be caused by a disturbed balance of the production and absorption of CSF leading to accumulation of CSF in the ventricular system or the subarachnoid space, or by impaired neurogenesis yielding abnormal amounts of CSF relative to the brain volume.[3]

Monogenic forms of HC exist,[4] but the genetic etiology is complex and > 100 genes associated with HC have been reported in the literature, however with highly variable penetrance. Interestingly, a nationwide population-based familial aggregation study of HC revealed preferential maternal inheritance.[2] The maternal component was not explained by risk factors likely to aggregate in families, [2] and may point towards X-linked genetic etiologies or involvement of mitochondrial DNA (mtDNA).

Many of the known HC-associated genes are involved in classical ciliopathies [5, 6] or in signaling pathways, such a sonic-hedgehog (Shh), wingless (Wnt), Notch, and PI3K-Akt-mTOR pathways, that are essential for brain development.[7] These pathways are compartmentalized - at least partly - in primary cilia.[8, 9] This role of ciliary function [10] and cilia-associated genes in HC is supported by the finding that genetically engineered mice with a HC phenotype overwhelmingly had genetic variants in cilia-associated genes.[11]

As ciliary function depends on a multitude of energy-requiring processes;[12] genetic or environmentally induced variation in OXPHOS function, or more broadly, mitochondrial function,[13] could modify the phenotype. This “Bioenergetic paradigm” [14] has been suggested to play a role in metabolic disorders/ biophysical traits,[15, 16] neurological diseases,[17] cardiomyopathies,[18, 19, 20, 21, 22] psychiatric diseases,[23] cancers,[24] and developmental disorders.[25] Furthermore, HC exhibits many comorbidities, including autism spectrum disease [26] that has been associated with altered mitochondrial function,[27, 28] and in some – but not all [29, 30] - studies with mtDNA haplogroups.[31] Mitochondrial DNA haplogroups, representing evolutionarily fixed groups of sequence variants in mtDNA, exhibit different characteristics of OXPHOS function [32, 33] and disease associations.[14, 34]

Based on the above, we hypothesized that variation in mtDNA haplogroups might explain the variable penetrance of HC. In consequence, we ascertained and compared mtDNA haplotypes and macro-haplotypes from HC cases and background population controls from the iPSYCH cohort.

## Materials and Methods

### Study design

This is a register-based case-cohort study using data from national health registries and genetic data from the iPSYCH study. The study is thus a sub-study of the Lundbeck Foundation Initiative for Integrative Psychiatric Research (iPSYCH: http://iPSYCH.au.dk). The iPSYCH cohort was selected from a study base of 1,472,762 singleton births born between May 1 1981 and Dec 31 2005. The inclusion criteria were that they were alive and residing in Denmark at one year of age, had an identifiable mother with a Danish Civil Registration number, and had a dried blood spot card stored in the Danish Neonatal Screening Biobank within the Danish National Biobank,[35]. The cohort comprised 57,875 psychiatric patients and a random sampled control cohort of 28,606 individuals. Details on time periods of selection as well as genomic variability data are given in Pedersen et al, 2018.[36]

### Ethics

The study was approved by the Scientific Ethics Committees of the Central Denmark Region (www.komite.rm.dk)(J.nr.: 1-10-72-287-12) and the Danish Data Protection Agency (www.datatilsynet.dk)(J.nr.: 2012-41-0110). Informed consent was waived in accordance with the conditions of ethics approval.[36]

### Hydrocephalus cases and controls

Hydrocephalus cases included individuals registered in the Danish National Patient Register 2016 or earlier with ICD 10 diagnosis code Q03, Q038, Q038A, Q038C, Q039, G910, G911, G912, G913, G918 and G919 or ICD 8 diagnosis code 74200, 74201, 74208, 7409, 34794 and 34795. A total of 191 hydrocephalus cases were identified in the iPSYCH cohort with haplotyped mtDNA and 24,831 background population controls. Among these, 172 hydrocephalus cases and 21,850 background population controls belonged to the European macro-haplogroups, HV, JT, and UK.

### Genetic analysis and mtDNA haplotyping

Two 3.2-mm disks were excised from each blood spot. DNA was extracted using Extract-N-Amp Blood PCR Kit (Sigma-Aldrich), whole genome amplified (WGA) in triplicate using the REPLIg kit (Qiagen), followed by pooling into a single aliquot. The amplified DNA samples were genotyped at the Broad Institute (MA, USA) using the PGC developed PsychChip v 1.0 (Illumina, CA, USA) typing 588,454 variants. Finally, samples with less than 97 % call rate, or where the estimated gender differed from the expected gender, were removed. We then isolated the 418 mitochondrial loci and reviewed the genotype calls, before exporting into the PED/MAP format using GenomeStudio (Illumina). MtDNA haplotyping was performed as described previously.[37, 38] using the haplogroup defining SNPs reported in www.phylotree.org.[39] Figure 1 illustrates the phylogenetic tree of mtDNA haplogroups.

**Figure 1.**
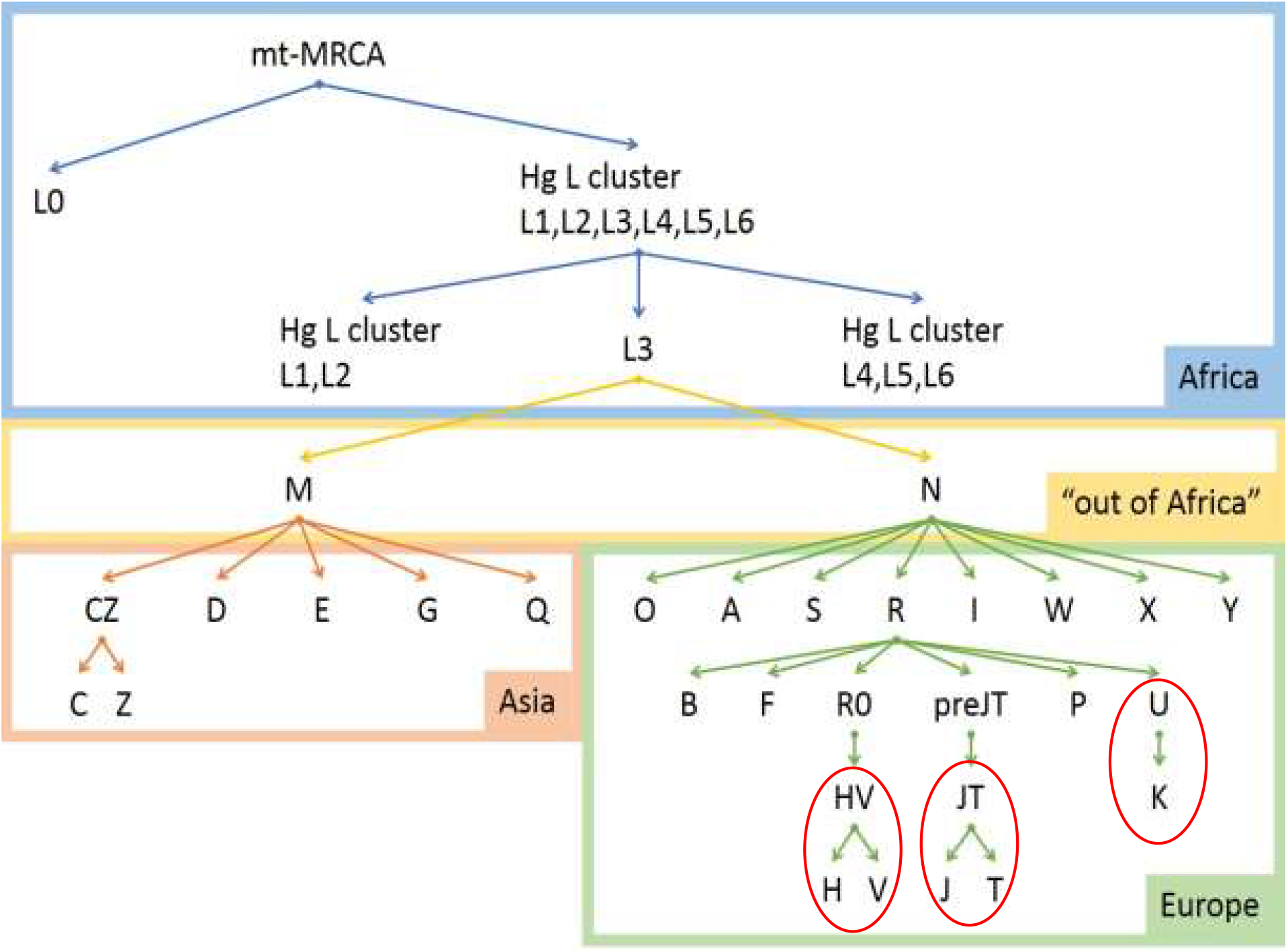
Phylogenetic tree of mtDNA haplogroups (www.phylotree.org). MRCA: Most recent common ancestor. The red circles denote the macro-haplogroups HV, JT, and UK.

### Statistics

The associations between HC and mtDNA haplogroups and macro-haplogroups (Figure 1) were examined using Fisher’s exact test. P-values less than 0.05 were considered significant.

## Results

We identified 191 cases of HC and 24,831 background population controls with available mtDNA. As presented in Table 1, 34 % of the HC cases were females, whereas 49 % of the background population controls were female. Mean age of HC cases was 22.4 years, which was virtually similar to the mean age of 22.8 years among background population controls.

**Table 1.** Demographics of the cohort of HC patients and controls.

|  | **HC cases** | **Controls** |
| --- | --- | --- |
| **Number** | 191 | 24,831 |
| **Sex (Women %)** | 34.6 | 49.0 |
| **Age, median (interquartile range)** | 22.4 (16.7 – 27.6) | 22.8 (17.5 – 28.7) |

Table 2 and Table 3 present the distribution of mtDNA haplogroups and mtDNA macro-haplogroups in HC cases and controls, respectively. The distribution of mtDNA haplogroups did not differ between HC cases and controls as shown in Table 2. Likewise, we observed no significant association between any of the European macro-haplogroups (HV, JT, and UK) and HC (Table 3).

**Table 2.**
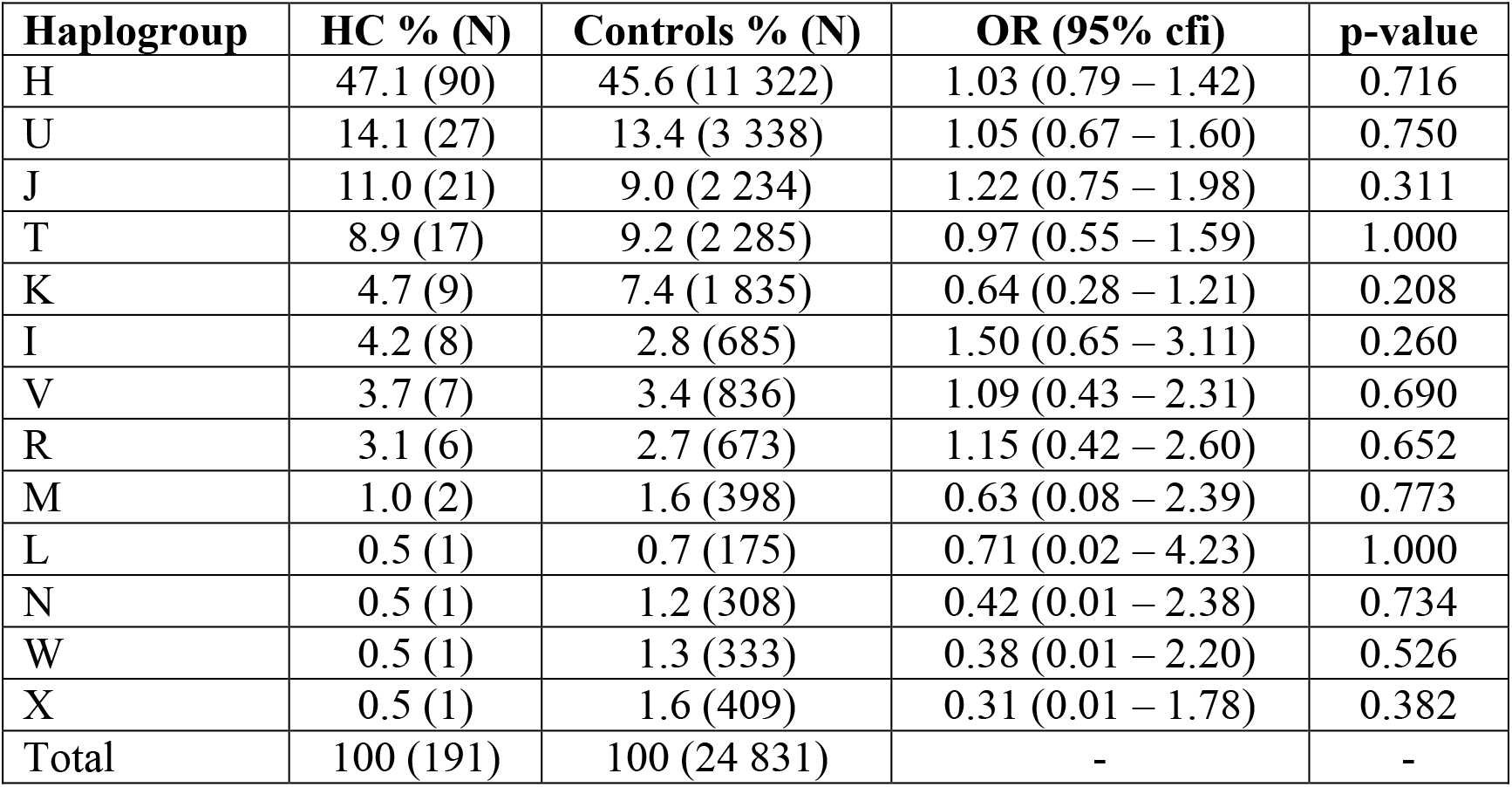
Distribution of mtDNA haplogroups in HC cases and controls.

| **Haplogroup** | **HC % (N)** | **Controls % (N)** | **OR (95% cfi)** | **p-value** |
| --- | --- | --- | --- | --- |
| H | 47.1 (90) | 45.6 (11 322) | 1.03 (0.79 – 1.42) | 0.716 |
| U | 14.1 (27) | 13.4 (3 338) | 1.05 (0.67 – 1.60) | 0.750 |
| J | 11.0 (21) | 9.0 (2 234) | 1.22 (0.75 – 1.98) | 0.311 |
| T | 8.9 (17) | 9.2 (2 285) | 0.97 (0.55 – 1.59) | 1.000 |
| K | 4.7 (9) | 7.4 (1 835) | 0.64 (0.28 – 1.21) | 0.208 |
| I | 4.2 (8) | 2.8 (685) | 1.50 (0.65 – 3.11) | 0.260 |
| V | 3.7 (7) | 3.4 (836) | 1.09 (0.43 – 2.31) | 0.690 |
| R | 3.1 (6) | 2.7 (673) | 1.15 (0.42 – 2.60) | 0.652 |
| M | 1.0 (2) | 1.6 (398) | 0.63 (0.08 – 2.39) | 0.773 |
| L | 0.5 (1) | 0.7 (175) | 0.71 (0.02 – 4.23) | 1.000 |
| N | 0.5 (1) | 1.2 (308) | 0.42 (0.01 – 2.38) | 0.734 |
| W | 0.5 (1) | 1.3 (333) | 0.38 (0.01 – 2.20) | 0.526 |
| X | 0.5 (1) | 1.6 (409) | 0.31 (0.01 – 1.78) | 0.382 |
| Total | 100 (191) | 100 (24 831) | - | - |

**Table 3.** Associations between macro-haplogroups and hydrocephalus.

| **Macro-haplogroup** | **HC % (N)** | **Controls % (N)** | **OR (95%-cfi)** | **p-value** |
| --- | --- | --- | --- | --- |
| HV | 56.3 (97) | 55.6 (12 158) | 1.03 (0.75 – 1.41) | 0.8777 |
| JT | 22.1 (38) | 20.7 (4 519) | 1.09 (0.74 – 1.57) | 0.6371 |
| UK | 21.5 (37) | 23.6 (5 173) | 0.88 (0.60 – 1.28) | 0.5888 |
| Total | 99.9 (172) | 99.9 (21 850) | - | - |

## Discussion

We have, for the first time, shown that genetic variation of mtDNA, at the level haplogroups and European macro-haplogroups is not associated with HC in the Danish population. This does not preclude that mtDNA variation at a more distant phylogenetic level, e.g. individual “private” variants or sub-haplogroups, may be of significance.

The mtDNA haplogroup variation is frequently found associated with late-presenting degenerative diseases, whereas rarer mtDNA variants often present as classical mitochondriopathies,[14, 34] which are maternally inherited. This rarer mtDNA model of disease would be more in line with the tendency for maternal inheritance found in the previously mentioned familial aggregation study[2]. Furthermore, rarer mtDNA diseases often negatively influence the ability to reproduce. Many rare mitochondrial diseases, either caused by mtDNA variants or nuclear genes coding for parts of the mitochondrial proteome or mitochondrial regulatory genes, have been described to present with HC, e.g. fatty acid oxidation defects,[40] mitochondrial respiratory complex deficiencies,[41, 42] loss of mitochondrial apoptosis-inducing factor,[43] and Fowler syndrome.[44]

Thus, this study does not exclude associations between rare mtDNA variants and hydrocephalus. Interestingly, we found hydrocephalus associated variants the *MTO1* gene in a single heterozygote carrier among a subset of 72 whole-exome sequenced hydrocephalus patients from the same cohort as this study (yet unpublished data). *MTO1* encodes the mitochondrial translocation optimization 1 protein, which is involved in tRNA modification to increase the accuracy and efficiency of mtDNA translation.[45] Furthermore, the study revealed significant involvement of the ciliome. Hydrocephalus has been associated with other mitochondrial gene mutations in experimental studies, either because of impaired brain development or loss of ciliated epithelium in the ependymal layer.[43]

A limitation of the study is that we have not examined the potential significance of variation in the nuclear genes coding for a large part (∼ 99 %) of the mitoproteome, and genes controlling interaction between mtDNA and the nuclear genome.[46] Such studies would have required a larger number of HC cases and much more extensive sequencing. Furthermore, the distribution of haplogroups in both controls and HC patients is similar to that previously reported for a large unbiased Danish nation-wide study [37] with such a great variation in frequency of different haplogroups (Table 2), that the study has insufficient statistical power to evaluate the association between the less frequent haplogroups and hydrocephalus.

In conclusion, we did not find significant associations between hydrocephalus and mtDNA at the haplogroup or macro-haplogroup levels. Yet, compelling evidence for the interaction between HC, brain development, ciliary-dependent signaling pathways,[47] and mitochondrial function exists,[13] but more likely due to rare mtDNA population variants. It has been shown that less frequent mtDNA variants are often mildly deleterious, and there have been attempts to link their collective impact with na number of phenotypes[48, 49, 50] We suggest that more extensive studies involving whole-exome-sequencing of the nuclear genome and full mtDNA sequencing are performed to better assess the role of mitochondrial function in modifying the complex genetic and molecular etiology of HC. This is of clinical importance as mitochondrial functional involvement may assist in defining new pharmaceutical targets.[51]

## Supporting information

Strobe checklist

## Data Availability

All data produced in the present study may be made available upon reasonable request to the authors and approval by the Danish Data Protection Agency, the iPSYCH PI group, and the Scientifics Ethics Committee of the Central Denmark Region.

## Acknowledgements

The iPSYCH study was funded by the Lundbeck Foundation Initiative for Integrative Psychiatric Research (www.ipsych.au.dk). This research has been conducted using the Danish National Biobank resource, supported by the Novo Nordisk Foundation.

## Declaration of interest statement

The authors report there are no competing interests to declare.

## Notes

### Competing Interest Statement

The authors have declared no competing interest.

### Funding Statement

The study did not receive any funding.

### Author Declarations

The study was approved by the Scientific Ethics Committees of the Central Denmark Region (www.komite.rm.dk)(J.nr.: 1-10-72-287-12) and the Danish Data Protection Agency (www.datatilsynet.dk)(J.nr.: 2012-41-0110). Informed consent was waived in accordance with the conditions of ethics approval

